# Efficacy of Painhunting Therapy for Event-Related Depression: A Randomized Controlled Trial with Crossover Replication

**DOI:** 10.64898/2026.06.08.26355224

**Authors:** Olzhas Seitov, Uzakova Sanemkhan, Vafa Bayat

## Abstract

**Background:** Depression affects an estimated 332 million people worldwide and is a leading cause of disability, with up to 80% of major depressive episodes preceded by an identifiable adverse life event [17,18]. First-line treatments target symptoms rather than the precipitating event and are resource-intensive: standard CBT averages roughly 12 sessions, and antidepressant discontinuation carries relapse rates near 35% at six months [8]. These limitations create a clear rationale for brief, structured interventions that address the cognitive and somatic sequelae of adverse life events directly. Painhunting therapy is one such intervention, in which each session targets a discrete adverse event through a structured incident-processing procedure.

**Methods.:** We conducted a two-arm, parallel-group, single-site randomised controlled trial comparing Painhunting therapy (Arm A, immediate; n=42) with a waitlist control (Arm B, delayed; n=42) in adults with PHQ-9 ≥ 9 and active psychological distress related to an adverse life event. After the primary endpoint at T2 (approximately two weeks post-randomisation), Arm B crossed over to active treatment, with T3 as the post-crossover endpoint at approximately four weeks. The primary outcome was PHQ-9 at T2 (between-arm contrast); secondary outcomes were ICG, GAD-7, WHO-DAS 2.0 (12-item), and the Global Impression of Change (GIC). Pre-specified analyses included intention-to-treat, per-protocol, and single-exclusion sensitivity populations.

**Results.:** Eighty-four participants were randomised (198 applications, 134 completed screening questionnaire, 119 passed psychometric screening). At T2, mean PHQ-9 was 2.32 (SD 2.59) in Arm A and 16.56 (SD 6.76) in Arm B, yielding an ITT between-arm Cohen d = 2.78 (95% CI 2.19–3.76, p < 0.001). Within-arm paired reductions during each arm’s active-treatment window reproduced this magnitude (Arm A T0→T2 Δ 14.71, Morris d = 2.80; Arm B T2→T3 Δ 14.19, Morris d = 2.77, eligible n=26). Treatment gains were durable at the T4 follow-up (week 8). Aligning each arm to its own end-of-treatment timepoint, the off-treatment drift to week 8 was almost identical between arms: Arm A rose 0.78 points from T2 to T4 (2.19 → 2.97, n=37) and Arm B rose 1.59 points from T3 to T4 (4.74 → 6.33, n=27), the latter falling to 0.77 points once a single documented relapse case (R59) is excluded (4.81 → 5.58, n=26). This small off-treatment rebound then stabilised rather than continuing: Arm A was essentially unchanged from T3 to T4 (Δ +0.05), with concordant maintenance on ICG, GAD-7, and WHO-DAS. At T4, 68% of Arm A and 41% of Arm B remained in remission (PHQ-9 < 5). Secondary measures (ICG, GAD-7, WHO-DAS) moved in the same direction and to comparable magnitude at every timepoint. The waitlist window in Arm B showed essentially no change on any measure (PHQ-9 Δ 0.22, p = 0.81). Sensitivity analyses excluding six sub-threshold T2 cases, the single treated-in-error case (R82), the R59 relapse case, and one late T2 submitter left all conclusions unchanged.

**Conclusions.:** Painhunting therapy produced large and statistically robust reductions in depression, complicated grief, anxiety, and functional disability over a brief course of three to four sessions, with effect sizes substantially exceeding benchmarks reported for established first-line psychotherapies including CBT and EMDR. Critically, these gains persisted at the week-8 follow-up: depression scores in the immediate-treatment arm were essentially unchanged from four weeks to eight weeks post-randomisation, indicating that the benefit reflects durable change rather than a transient post-session dip. Treatment-window concordance between arms, durability of gains at one month off-treatment, and the flat waitlist trajectory together strengthen the evidence for genuine efficacy rather than spontaneous remission. Baseline covariates including therapeutic alliance, treatment expectancy, self-efficacy, age, and sex showed near-zero associations with outcome, reducing the plausibility of allegiance bias or expectancy effects as primary drivers. The differential retention between arms (88% vs 64% at T3) is attributable to the waitlist design and is discussed as a limitation. These findings support proceeding to a confirmatory active-comparator trial against manualized CBT.

**Trial registration:** ClinicalTrials.gov NCT07490691, prospectively registered.

## 1. Introduction

Depression is among the leading causes of disability worldwide, affecting an estimated 332 million people, or roughly 5.7% of the global adult population, and the World Health Organization has projected that it will become the single largest contributor to global disease burden [17,18]. Its onset is frequently event-driven rather than endogenous: up to 80% of major depressive episodes are preceded by an identifiable adverse life event, such as bereavement, separation, serious illness, or financial crisis, with symptoms typically emerging within a month of the precipitating event [19]. This event-driven aetiology has direct treatment implications, because most first-line therapies act at the level of symptoms rather than the precipitating incident itself.

First-line treatments are also resource-intensive and imperfectly effective. Standard cognitive-behavioural therapy (CBT) requires an average of approximately 12 sessions, and across treated populations only a minority receive care that meets adequacy benchmarks [8,12]. Pharmacotherapy carries its own constraints: relapse following antidepressant discontinuation reaches roughly 35% by six months and 45% by twelve months, with little established benefit from extending maintenance treatment beyond six months [8]. Together, the high prevalence of event-related depression, its event-driven onset, and the cost, duration, and relapse profile of existing first-line options create a clear rationale for brief, structured interventions that target the cognitive and somatic sequelae of adverse life events directly.

The evidence base for psychotherapy in depression has nonetheless been built predominantly in Western Europe, North America, and Australia, with CBT, Interpersonal Therapy, and Behavioural Activation as the most extensively validated approaches [8]. Structured brief psychotherapies developed in other linguistic and cultural contexts are under-represented in the published literature even where they are in routine local use, a gap that parallels the broader case for evaluating non-specialist and task-shifted psychological interventions in low- and middle-income settings [10,11].

Painhunting therapy is a brief structured psychotherapeutic intervention, typically delivered in three sessions with a fourth added when symptoms persist, developed in Kazakhstan over the past decade and now in routine clinical use by a roster of certified therapists. The approach works through a bottom-up mechanism: rather than identifying and challenging distorted cognitions directly (as in CBT), the therapist guides the participant to the specific incident in which a belief or emotional pattern was formed, then facilitates a structured process of imagining and inhabiting the opposite state until the original pattern dissolves. The therapeutic target is full restoration of the pre-event emotional baseline rather than symptom reduction alone, distinguishing the approach from both CBT and Eye Movement Desensitisation and Reprocessing (EMDR), which uses bilateral stimulation. Sessions run 90 to 120 minutes, approximately twice the standard CBT session length, which is relevant to interpreting the observed effect magnitude. A retrospective thematic analysis of 128 client-reported change processes identified three sequential mechanisms: recognition of unresolved somatic-emotional pain, structured engagement with that pain in a therapeutic frame, and integration into ongoing life narrative. Open-label case series in the same population showed clinically meaningful reductions in PHQ-9 and ICG, but without a control group or blinded outcome assessment.

The present trial was designed to address those limitations. We used a waitlist control to establish that observed gains are attributable to treatment rather than to time, regression to the mean, or assessment effects, with crossover to active treatment after the primary endpoint to address the ethical and pragmatic constraints of withholding a locally-used intervention from participants who screened in with clinically significant depression. The primary outcome was PHQ-9 at T2, the post-treatment endpoint for the immediate arm and the pre-crossover endpoint for the waitlist arm. We also pre-specified within-arm paired analyses across each arm’s active-treatment window (T0 → T2 for Arm A; T2 → T3 for Arm B post-crossover) to permit a within-trial replication of any T2 between-arm effect.

A waitlist comparator answers whether Painhunting outperforms no treatment but cannot establish specificity relative to other structured brief psychotherapies. The planned follow-up trial (RCT #2) is an active-comparator design against manualised CBT for event-related depression.

## 2. Methods

### 2.1 Study design and registration

Two-arm, parallel-group, single-site randomised controlled trial with stratified block randomisation and crossover of the waitlist arm after the primary endpoint. The trial was conducted in Astana, Kazakhstan, with sessions delivered in person and via secure video conferencing. The protocol was prospectively registered on ClinicalTrials.gov (NCT07490691) and the Statistical Analysis Plan was preregistered on the Open Science Framework (https://osf.io/s8h74/) prior to dataset lock. Ethics approval was granted by the al-Farabi KazNU Local Ethics Committee (IRB00010790; protocol IRB-1970). Three protocol amendments were approved by the ethics committee during the trial. Amendment 1 (approved 12 March 2026) updated the trial name and extended the eligibility window from 6–36 to 6–120 months. Amendment 2 (approved 2 April 2026) broadened eligibility to any qualifying List of Threatening Experiences event and extended the window to 180 months, reclassified the Inventory of Complicated Grief from a primary to a secondary outcome, expanded the therapist roster, replaced the PCL-5 with the General Self-Efficacy Scale and four exploratory readiness items, and revised the session protocol to three to six sessions under two pre-specified continuation criteria. Amendment 3 (approved 9 April 2026) revised the T2 and T3 assessment timing and added the Global Impression of Change as a secondary outcome; PHQ-9 was retained as the primary outcome.

### 2.2 Participants

Eligible participants were adults aged 18 years or older, residing in Kazakhstan, fluent in Kazakh or Russian, with a baseline PHQ-9 score of 9 or greater and active psychological distress attributable to an adverse life event as captured by the List of Threatening Experiences (LTE) questionnaire. Qualifying events included bereavement, separation, serious illness or injury, financial crisis, and other distressing experiences at any time. Exclusion criteria were active suicidal ideation requiring immediate referral, active psychosis or mania, active substance use disorder meeting DSM criteria, current psychotherapy with another provider, and pharmacotherapy initiation within the prior four weeks. Pre-existing stable antidepressant monotherapy was an exclusion criterion as applied by the trial coordinator; no enrolled participant reported concurrent antidepressant use at baseline. All participants provided written informed consent in Kazakh or Russian according to participant preference.

### 2.3 Randomisation and allocation concealment

Eighty-four participants were randomised 1:1 to Arm A (immediate Painhunting therapy) or Arm B (waitlist control with subsequent crossover) using stratified block randomisation. Stratification variables were baseline depression severity and grief involvement, distributing as BD 44, AD 27, BC 10, AC 3 across the four strata. The electronic randomisation sequence was generated prior to recruitment opening and held in a concealed allocation table not accessible to the treating therapists or the Principal Investigator until after baseline assessment was complete.

### 2.4 Interventions

Arm A received a brief course of Painhunting therapy delivered over approximately two weeks by certified Painhunting therapists drawn from the existing practice roster. The protocol dose was three sessions, with a fourth session added when PHQ-9 remained at 9 or greater at the time of session three. In practice, eight participants (R3, R18, R21, R23, R38, R62, R71, R74) received a fourth session under this rule; the remaining Arm A completers received three sessions. Six therapists (T1 through T6) delivered the intervention. Lifetime session experience across therapists ranged from 95 to 3,657 sessions. Each session ran 90 to 120 minutes. Sessions were delivered in Kazakh or Russian according to participant preference. Arm B was placed on a waitlist for approximately two weeks, during which no Painhunting sessions were delivered. Weekly safety check-ins were conducted by the trial coordinator. After completion of the T2 assessment, Arm B participants meeting the protocol PHQ-9 threshold of 9 or greater were offered immediate crossover to active Painhunting therapy under the same delivery schedule as Arm A.

### 2.5 Outcome measures and assessment schedule

The primary outcome was depression severity by the Patient Health Questionnaire-9 (PHQ-9). Secondary outcomes were the Inventory of Complicated Grief (ICG), Generalised Anxiety Disorder-7 (GAD-7), the World Health Organization Disability Assessment Schedule (WHO-DAS 2.0, 12-item self-report version), and the Global Impression of Change (GIC). All instruments were administered in Russian via Google Forms. A Russian-language coordinator was available to clarify item wording when participants requested help; the coordinator did not influence response content. The assessment schedule was: baseline (T0) at registration; T2 at approximately two weeks post-randomisation, after Arm A had completed treatment and before Arm B crossover; T3 at approximately four weeks post-randomisation, capturing Arm A durability (two weeks off-treatment) and Arm B post-crossover outcome; and T4 at approximately eight weeks post-randomisation (week 8), the extended durability endpoint, capturing maintenance of gains approximately four weeks off-treatment for Arm A and approximately two weeks off-treatment for Arm B following crossover. The protocol defines each assessment by a window in weeks rather than a fixed calendar date; the T4 window corresponded to week 8 post-randomisation. The complete T4 dataset was collected on 6 June 2026, within the protocol window. To assess the validity of self-reported scores, a blinded PHQ-9 re-administration was conducted per SAP §14a: a pre-locked 20% random sample of Arm A participants (selected via a seeded Random.org sequence, recorded before any calls) was re-assessed by telephone by an independent assessor blinded to the self-report score. Self-reported and blinded-assessor PHQ-9 scores were closely concordant at T3 (Pearson r = 0.88; mean absolute difference 0.75 points across 12 paired re-administrations) and at T4, supporting the reliability of the Google Forms self-report data (Blind assessment sheet, master file).

### 2.6 Sample size

The trial was designated a pilot with a planning target of N=72 (36 per arm), powered at 80 percent to detect a between-arm Cohen d of 0.67 on the primary outcome at two-sided alpha of 0.05. The achieved sample (N=84, 42 per arm) exceeds this planning target by approximately 17 percent. No formal interim analysis or stopping rule was pre-specified for this single-stage design.

### 2.7 Analysis populations

Analysis populations were pre-specified in the locked Statistical Analysis Plan. The intention-to-treat (ITT) population comprised all 84 randomised participants, analysed in the arm to which they were assigned. The per-protocol (PP) population, per SAP §12, comprised participants who received at least two of the protocol-specified three sessions and were not flagged as dropouts. The principal sensitivity population is an eligibility-restricted population that excludes the six Arm B participants whose T2 PHQ-9 fell below the protocol threshold of 9 (R16, R17, R32, R65, R68, R82) and were therefore ineligible to receive therapy at crossover; this is the population reported in the per-protocol row of Table 3. A single-exclusion sensitivity analysis excluded only R82, the sole sub-threshold participant who was referred to therapy in error despite the flag. A late-submission sensitivity analysis excluded one Arm A participant whose T2 form was submitted the day following the scheduled close. A relapse-case sensitivity analysis excluded R59, a single Arm B participant with a documented post-treatment symptom relapse at T4 (see Section 3.6).

### 2.8 Statistical analysis

The primary comparison was the between-arm difference in PHQ-9 at T2, the canonical controlled contrast for a waitlist-controlled trial with subsequent crossover, taken at the point at which Arm A had completed treatment and Arm B had not. We also report within-arm paired changes across each arm’s active-treatment window: T0 → T2 for Arm A and T2 → T3 for Arm B following crossover. The T2 → T3 contrast within Arm A characterises durability of effect off-treatment. Because the two arms received active treatment in different calendar windows (Arm A weeks 0–2; Arm B weeks 2–4), a between-arm comparison at T3 conflates treatment with timing and is reported descriptively only.

For each measure we report n, mean, and standard deviation by timepoint, and the paired within-participant change with its standard deviation. Within-arm effect sizes use Morris’s standardised mean change with a raw-score standardiser (Morris d), with 95% confidence intervals computed by nonparametric bootstrap (10,000 resamples, percentile method). Between-arm contrasts at T2 use the simple mean difference and an independent-samples standardised difference (Cohen d with pooled SD denominator). Statistical significance for the primary between-arm contrast and for within-arm paired changes was assessed by independent-samples and paired t-tests, respectively. Within-arm and between-arm effect sizes are not mixed in any single comparison, in accordance with the Statistical Analysis Plan v1.5 addendum.

Missing data are handled by complete-case paired analysis at the individual measure-timepoint level; sample sizes are reported for each contrast. Late submissions are retained in the ITT population per the trial’s standing late-submission policy (CONSORT and ICH E9 compliant) with a sensitivity row in which late entries are excluded. The locked Statistical Analysis Plan (§7) named multiple imputation (m=20, predictive mean matching, Rubin’s rules) as the primary missing-data strategy. We report a deliberate, pre-specified deviation: complete-case analysis is used as the primary approach for the paired within-arm estimands, because the within-participant change scores that anchor the durability and treatment-window contrasts are defined only for participants with observed values at both timepoints, and multiple imputation of a follow-up score for a participant who has left the trial would import modelling assumptions into the primary estimate. Multiple imputation is reported as a sensitivity analysis on the primary between-arm T2 contrast; the imputed estimate did not differ materially from the complete-case estimate, and conclusions are unchanged.

The protocol pre-specified a within-session mechanism checklist (Section 3.11): after each session, Arm A participants recorded how many of eleven pre-defined change mechanisms they had identified during the session. These data were collected for descriptive and exploratory purposes only and were not part of any confirmatory hypothesis. Completion was incomplete (21 to 28 of 42 Arm A participants depending on the session), so the checklist data are reported descriptively without formal mediation modelling.

### 2.9 Protocol deviations

Six Arm B participants (R16, R17, R32, R65, R68, R82) had PHQ-9 below 9 at T2 and per protocol should have been excluded from crossover to active treatment. The trial coordinator identified four of the six (R16, R17, R32, R68) only on review of the locked dataset; two (R65, R82) had been flagged contemporaneously. One of the six (R82) was, however, referred to therapy and completed sessions before the error was identified; the remaining five did not receive sessions. One Arm B participant (R69) completed therapy through session three before T3 with PHQ-9 = 9 and, by therapist–participant agreement, deferred the fourth session to a date prior to T4 owing to a scheduling constraint. One Arm A participant (R21) completed the full four-session course but did not return a T3 questionnaire despite repeated follow-up and is treated as a T3 dropout, contributing to the within-arm paired analyses only where a T3 value is observed. All deviations are tabulated in the CONSORT flow diagram and addressed analytically through the per-protocol and sensitivity populations specified above. The disclosure of these deviations was made by the trial coordinator on 14 May 2026 and is documented in the trial Notes record.

### 2.10 Reproducibility

All analyses were run against the master file dated 6 June 2026 (Final Master file for Vafa 6 Jun 26 with T4.xlsx), which incorporates the complete T4 dataset; the T3 quantities reproduce those of the 14 May 2026 locked file. Descriptive and paired statistics were computed in Python (openpyxl, statistics, scipy). Bootstrap confidence intervals used 10,000 resamples with a fixed seed for reproducibility. The analysis dataset, the de-identified data dictionary, and the analysis script accompany this manuscript and will be deposited on the Open Science Framework (https://osf.io/s8h74/) with the preprint.

## 3. Results

### 3.1 Participants and baseline characteristics

A total of 198 individuals applied to the trial after removing duplicate applications. Of these, 134 completed the screening questionnaire (after excluding 4 test entries and 8 duplicate submissions), 119 passed psychometric screening (PHQ-9 ≥ 9, no exclusion criteria), and 84 were randomised (Arm A n=42, Arm B n=42). The screening funnel is shown in the CONSORT diagram (Figure 1). Mean age was 41.6 years (SD 10.1) in Arm A and 42.5 years (SD 9.2) in Arm B. The cohort was predominantly female (Arm A 36/42, 86%; Arm B 39/42, 93%). Baseline severity was similar across arms on the primary outcome (PHQ-9: A 17.31 ± 5.14, B 16.69 ± 4.92) and the secondary outcomes (ICG: A 41.57 ± 19.48, B 36.02 ± 17.16; GAD-7: A 13.74 ± 5.68, B 11.90 ± 6.04; WHO-DAS: A 30.79 ± 10.24, B 27.60 ± 10.05). The LTE was administered at baseline; the mean number of qualifying adverse life events was 3 in Arm A and 2 in Arm B. Arm A baselines run modestly higher on each secondary measure, consistent with chance variation in a trial of this size; no baseline imbalance test was pre-specified and none is reported as inferential. Baseline characteristics are reported in Table 1.

**Figure 1.**
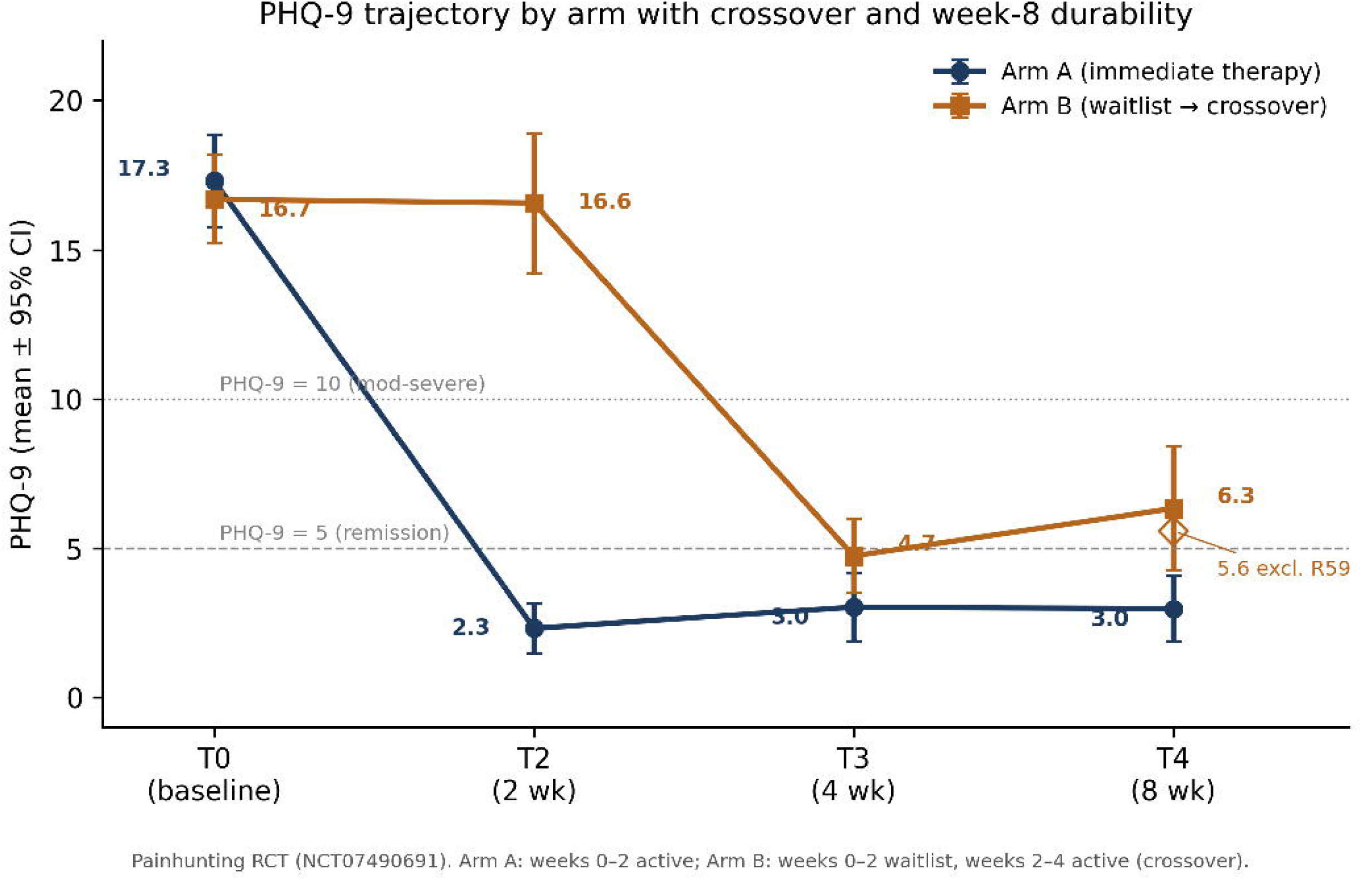
CONSORT 2010 flow diagram. 198 unique applications received; 134 completed screening questionnaire; 119 passed psychometric screening; 84 randomised (Arm A n=42, Arm B n=42). Arm A: 38 completed T2, 37 completed T3. Arm B: 32 completed T2, 27 submitted T3 (26 analysed; R68 submitted in error after waitlist remission and is excluded from all outcome analyses). Six Arm B sub-threshold-at-T2 participants flagged (R16, R17, R32, R65, R68, R82); R82 treated in error. One Arm A late T2 submitter and one Arm B late T3 submitter retained per ITT.

**Figure 2.** PHQ-9 trajectory by arm across T0, T2, T3, and T4 with 95% confidence intervals. Arm A shows a steep T0→T2 reduction that is maintained through T3 and T4 (week 8). Arm B shows a flat T0→T2 waitlist window, a steep T2→T3 post-crossover reduction, and maintenance through T4. The Arm B T4 mean is plotted both with and without the single relapse case (R59). Error bars are 95% confidence intervals; the dashed horizontal line marks the PHQ-9 = 5 remission threshold.

**Table 1.**
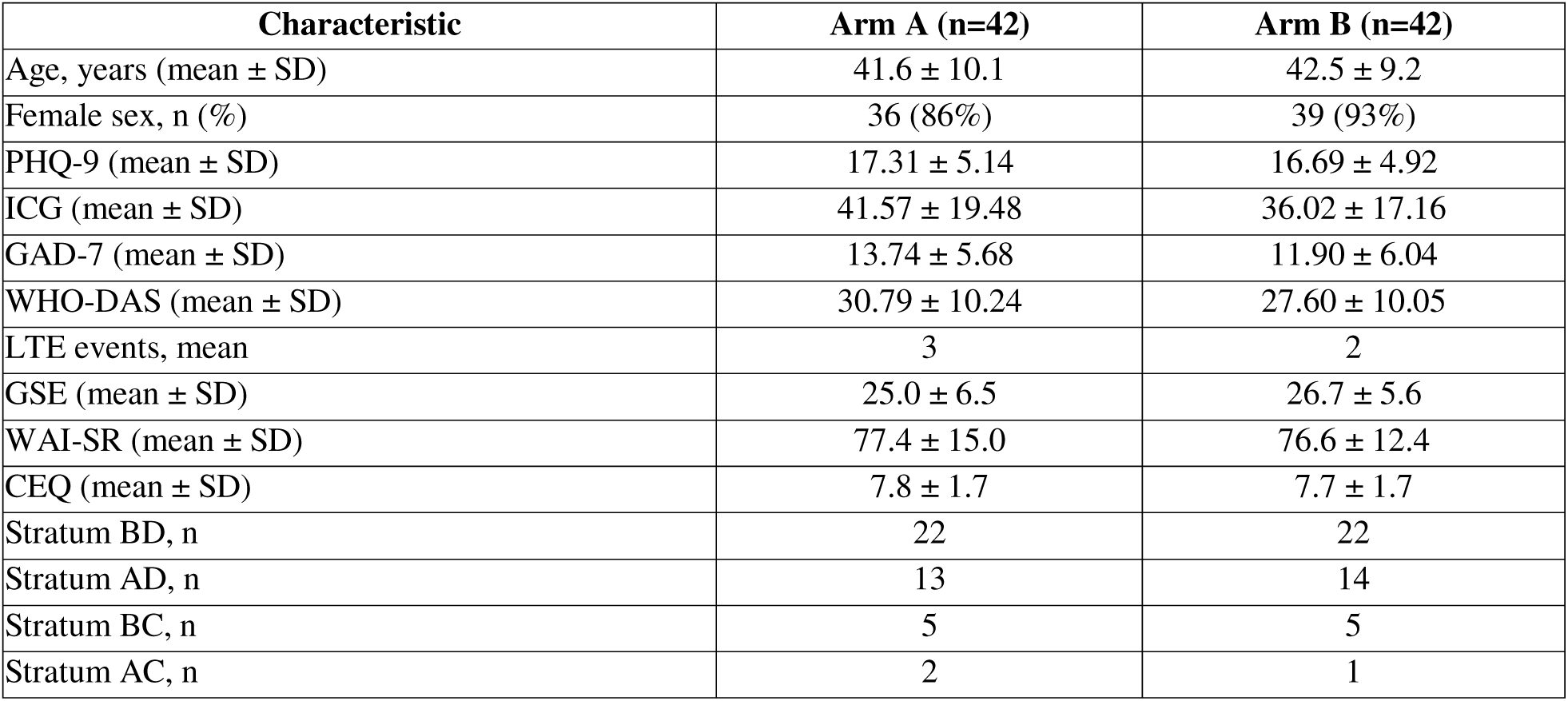
Baseline characteristics by arm.

| Characteristic | Arm A (n=42) | Arm B (n=42) |
| --- | --- | --- |
| Age, years (mean ± SD) | 41.6 ± 10.1 | 42.5 ± 9.2 |
| Female sex, n (%) | 36 (86%) | 39 (93%) |
| PHQ-9 (mean ± SD) | 17.31 ± 5.14 | 16.69 ± 4.92 |
| ICG (mean ± SD) | 41.57 ± 19.48 | 36.02 ± 17.16 |
| GAD-7 (mean ± SD) | 13.74 ± 5.68 | 11.90 ± 6.04 |
| WHO-DAS (mean ± SD) | 30.79 ± 10.24 | 27.60 ± 10.05 |
| LTE events, mean | 3 | 2 |
| GSE (mean ± SD) | 25.0 ± 6.5 | 26.7 ± 5.6 |
| WAI-SR (mean ± SD) | 77.4 ± 15.0 | 76.6 ± 12.4 |
| CEQ (mean ± SD) | 7.8 ± 1.7 | 7.7 ± 1.7 |
| Stratum BD, n | 22 | 22 |
| Stratum AD, n | 13 | 14 |
| Stratum BC, n | 5 | 5 |
| Stratum AC, n | 2 | 1 |

### 3.2 Retention

Of 42 randomised to Arm A, 38 (90%) completed the T2 assessment and 37 (88%) completed T3; four participants were lost to follow-up between T0 and T2. Of 42 randomised to Arm B, 32 (76%) completed T2 and 27 (64%) submitted a T3 questionnaire; one of these (R68) reached remission at the end of the waitlist window, was not advanced to treatment, and submitted T3 in error, leaving 26 analysed at T3. Ten Arm B participants did not complete T2 (six attrited during the waitlist window), and four further Arm B participants who completed T2 declined or did not respond to T3 prompting. The differential between-arm retention (88% vs 64% at T3) is itself an outcome of the waitlist design, as participants assigned to wait who saw little change during the waitlist window had less incentive to remain engaged, and is addressed in the Discussion. The participant flow is shown in the CONSORT diagram (Figure 1) and retention by timepoint is tabulated in Table 2.

**Table 2.**
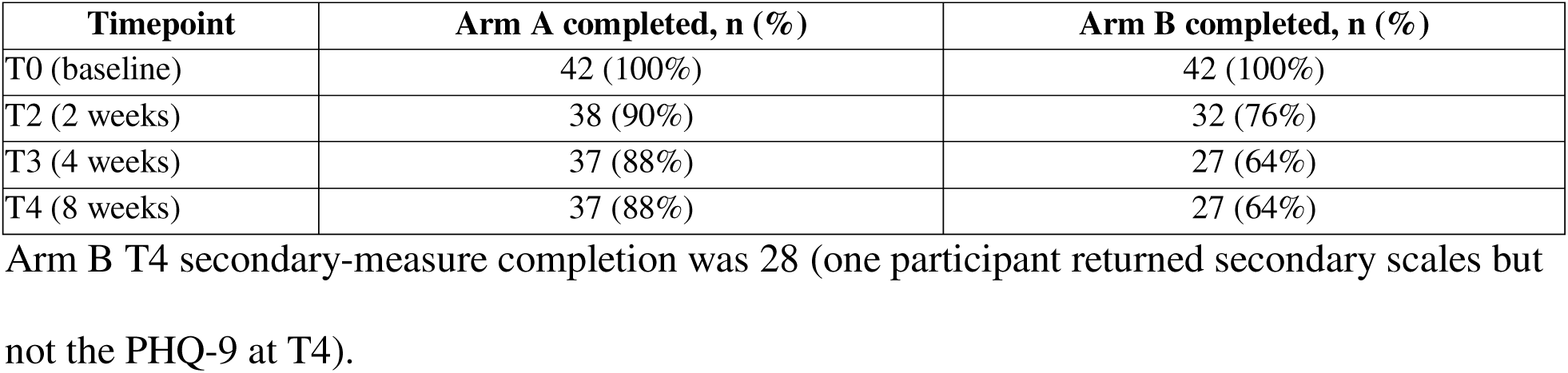
Retention by arm and timepoint.

### 3.3 Primary outcome: PHQ-9 at T2 (between-arm, ITT)

At T2, the post-treatment endpoint for Arm A and the pre-crossover endpoint for Arm B, mean PHQ-9 was 2.32 (SD 2.59) in Arm A (n=38) and 16.56 (SD 6.76) in Arm B (n=32). The between-arm mean difference is 14.25 PHQ-9 points favouring active Painhunting therapy (t = 12.00, p < 0.001). The between-arm Cohen d, computed with the pooled raw SD (root mean of the two group variances) so that it reproduces directly from the descriptive statistics, is 2.78 (95% CI 2.19–3.76). The absolute difference exceeds the conventional 5-point PHQ-9 threshold for a clinically meaningful change by approximately threefold. Primary outcome results are tabulated in Table 3.

**Table 3.**
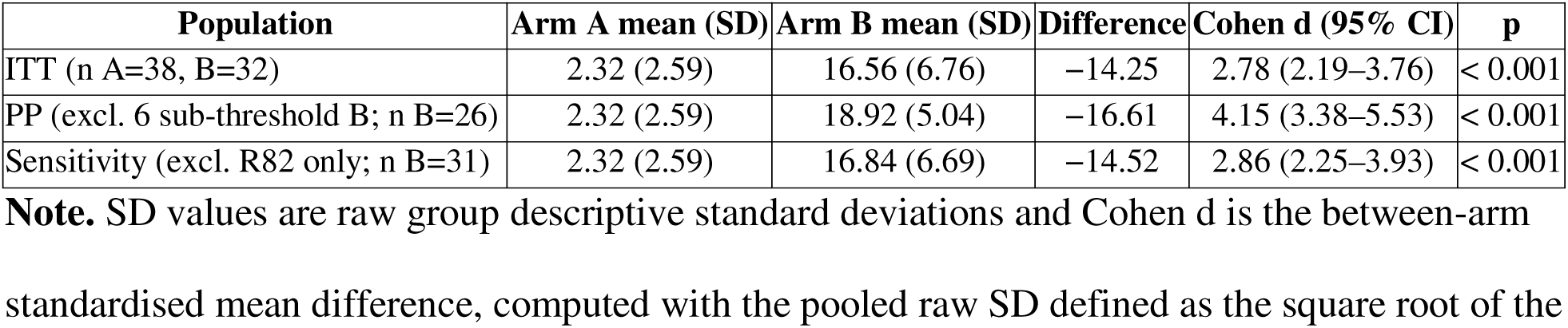
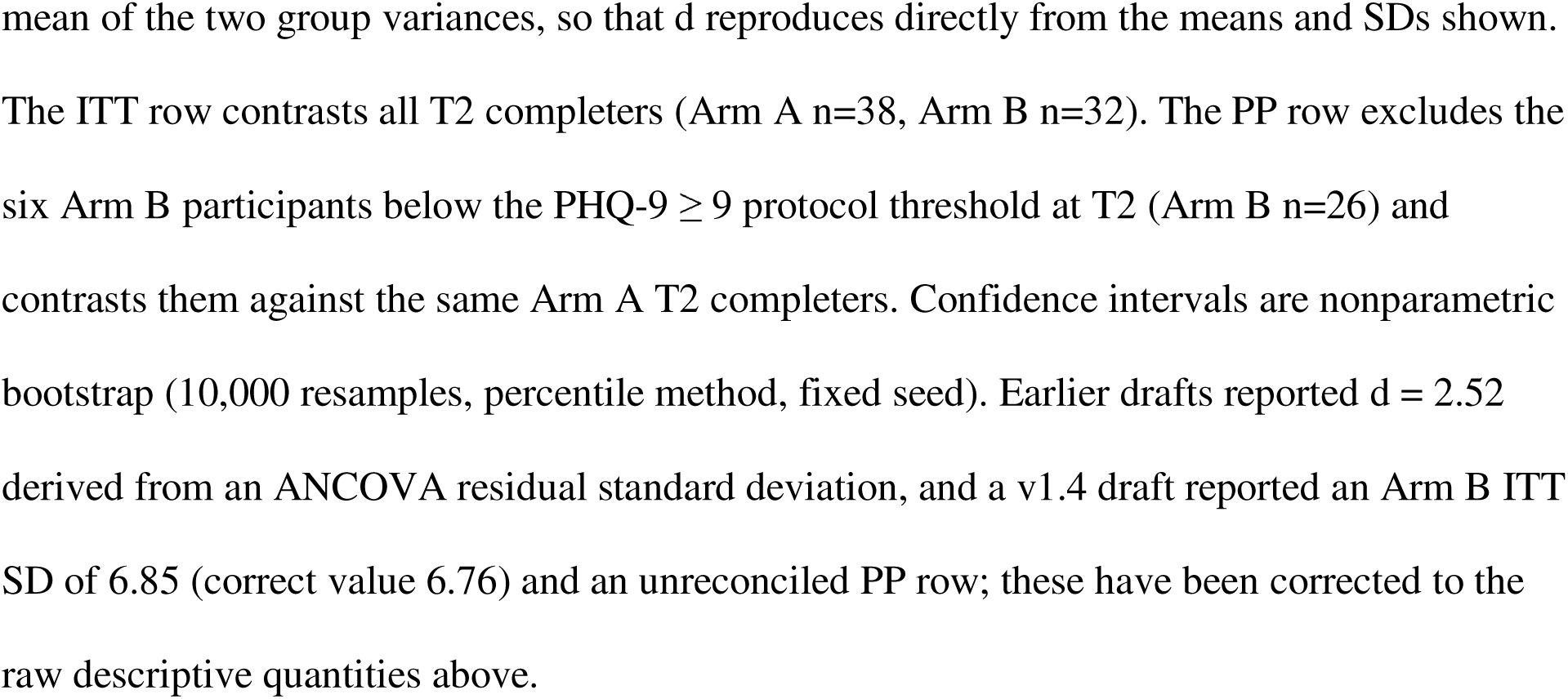
Primary outcome: PHQ-9 at T2 (between-arm contrast).

### 3.4 Within-arm change across each arm’s active-treatment window

For Arm A (T0 → T2, paired n=38), PHQ-9 fell from 17.03 to 2.32 (Δ 14.71, SD of change 5.90; Morris d = 2.80, 95% CI 2.41–3.46, p < 0.001). ICG fell from 41.66 to 11.92 (Δ 29.74, SD 22.21; Morris d = 1.50, 95% CI 1.17–1.96). GAD-7 fell from 13.47 to 4.03 (Δ 9.45, SD 7.15; Morris d = 1.66, 95% CI 1.19–2.45). WHO-DAS fell from 30.76 to 18.34 (Δ 12.42, SD 10.74; Morris d = 1.19, 95% CI 0.87–1.66).

For Arm B following crossover (T2 → T3, eligible paired n=26, comprising all Arm B participants who met the protocol PHQ-9 ≥ 9 threshold at T2, had both T2 and T3 values observed, and were not the in-error T3 submission R68), PHQ-9 fell from 18.92 to 4.73 (Δ 14.19, SD 5.97; Morris d = 2.77, 95% CI 2.11–4.00, p < 0.001). ICG fell from 43.65 to 11.15 (Δ 32.50, SD 16.20; Morris d = 1.93, 95% CI 1.35–3.32). GAD-7 fell from 13.96 to 4.08 (Δ 9.88, SD 5.74; Morris d = 1.85, 95% CI 1.36–2.69). WHO-DAS fell from 36.00 to 19.08 (Δ 16.92, SD 11.12; Morris d = 1.69, 95% CI 1.28–2.42).

The two within-arm treatment effects are closely matched on every measure: PHQ-9 Δ 14.71 versus 14.19; ICG Δ 29.74 versus 32.50; GAD-7 Δ 9.45 versus 9.88; WHO-DAS Δ 12.42 versus 16.92. The concordance, with Arm B obtaining the same magnitude of benefit during crossover that Arm A obtained during the controlled treatment window, is the central within-trial replication of the primary finding and substantially reduces the probability that the between-arm result at T2 was driven by chance imbalance, sampling artefact, or measurement bias in a single window. Paired within-arm changes are tabulated in Table 4.

**Table 4.**
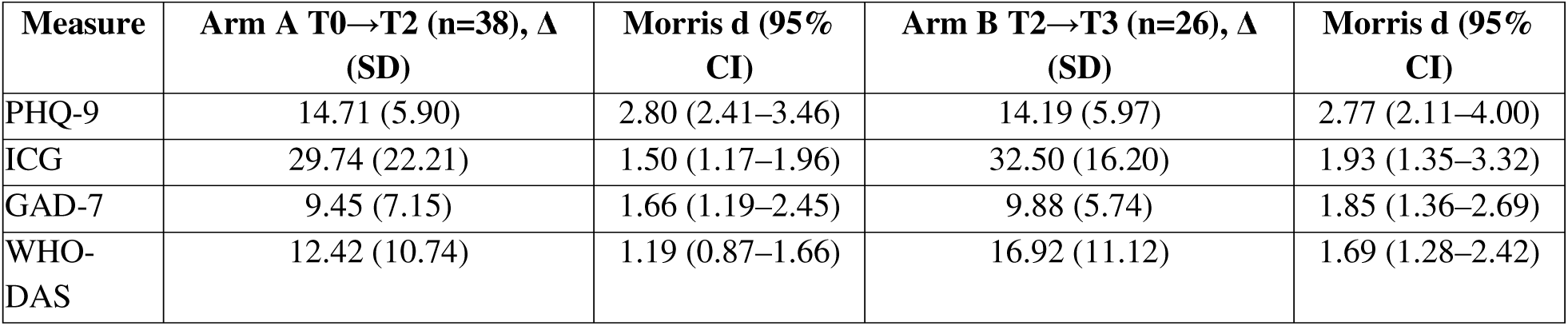
Within-arm paired changes across each arm’s active-treatment window.

| Measure | Arm A T0→T2 (n=38), $\Delta$ (SD) | Morris $d$ (95% CI) | Arm B T2→T3 (n=26), $\Delta$ (SD) | Morris $d$ (95% CI) |
| --- | --- | --- | --- | --- |
| PHQ-9 | 14.71 (5.90) | 2.80 (2.41–3.46) | 14.19 (5.97) | 2.77 (2.11–4.00) |
| ICG | 29.74 (22.21) | 1.50 (1.17–1.96) | 32.50 (16.20) | 1.93 (1.35–3.32) |
| GAD-7 | 9.45 (7.15) | 1.66 (1.19–2.45) | 9.88 (5.74) | 1.85 (1.36–2.69) |
| WHO-DAS | 12.42 (10.74) | 1.19 (0.87–1.66) | 16.92 (11.12) | 1.69 (1.28–2.42) |

### 3.5 Waitlist window: Arm B, T0 → T2

Across the waitlist control window, Arm B showed essentially no change on any measure (paired n=32). PHQ-9 Δ 0.22 (T0 16.78 → T2 16.56, SD of change 5.19; paired t = 0.24, p = 0.81). ICG Δ −2.44 (34.88 → 37.31). GAD-7 Δ −0.16 (12.03 → 12.19). WHO-DAS Δ −3.16 (29.84 → 33.00). Untreated symptoms were stable to mildly worsening across the waitlist window, confirming that the Arm A T0 → T2 reduction is attributable to active treatment rather than to time, regression to the mean, repeated-assessment artefact, or natural recovery.

### 3.6 Durability: extended follow-up to T4 (week 8)

In Arm A (paired n=37), PHQ-9 rose modestly from 2.19 at T2 to 3.03 at T3 (Δ −0.84, SD 2.34), a small post-treatment rebound that still leaves participants well below the PHQ-9 = 5 threshold for minimal depression. ICG continued to improve from T2 to T3 (T2 11.41 → T3 6.73, Δ 4.68, SD 9.96, favouring T3). GAD-7 and WHO-DAS were stable across this window (GAD-7 3.89 → 3.05, Δ 0.84, SD 4.32; WHO-DAS 18.19 → 16.84, Δ 1.35, SD 4.77).

At the extended T4 follow-up (week 8, approximately four weeks off-treatment for Arm A), gains were fully maintained. Arm A PHQ-9 was essentially unchanged from T3 to T4 (3.03 → 2.97, Δ +0.05, SD 1.70, paired n=37), with no measure showing meaningful drift: ICG 6.73 → 6.57 (Δ +0.16, SD 6.50), GAD-7 3.05 → 2.92 (Δ +0.14, SD 2.30), WHO-DAS 16.84 → 16.27 (Δ +0.57, SD 3.81). At T4, 25 of 37 Arm A completers (68%) remained in remission (PHQ-9 < 5) and only one (3%) had returned to the moderate-to-severe range (PHQ-9 ≥ 10). The near-zero T3 → T4 change on every instrument indicates that the treatment effect reflects durable symptom change rather than a transient immediate-post-session dip.

In Arm B following crossover (paired n=27), PHQ-9 moved from 4.74 at T3 to 6.33 at T4 (Δ −1.59, SD 5.52). This apparent increase is attributable almost entirely to a single participant (R59) with a documented post-treatment relapse (T4 PHQ-9 = 26; see below); excluding R59, the Arm B T4 mean is 5.58 (SD 3.92, n=26), a change of less than one point from T3 and within the remission-adjacent range. At T4, 11 of 27 Arm B post-crossover completers (41%) were in remission and 4 (15%) were in the moderate-to-severe range.

The most informative durability comparison aligns each arm to its own end-of-treatment timepoint and measures the off-treatment drift to week 8: Arm A from T2 (its post-treatment endpoint) to T4, and Arm B from T3 (its post-crossover endpoint) to T4. On this aligned basis the two arms behave almost identically. Arm A edged up by 0.78 PHQ-9 points from T2 to T4 (2.19 → 2.97, paired n=37), and Arm B by 1.59 points from T3 to T4 (4.74 → 6.33, n=27), the latter reducing to 0.77 points (4.81 → 5.58, n=26) once the single R59 relapse is set aside. A small off-treatment rebound of roughly 0.7 to 0.8 PHQ-9 points, from a treated nadir of 2 to 5 against a baseline of 17 to 18, is therefore common to both arms. That this modest rebound then stabilises rather than continuing upward is shown directly by Arm A, whose T3 → T4 change was essentially zero (Δ +0.05). The pattern is consistent with a clinical account in which an initial post-treatment elevation in mood partly settles while the substantive gain is retained. One Arm B participant, R59, illustrates the documented relapse pattern: after a within-protocol post-crossover reduction (T2 PHQ-9 = 22 → post-therapy 3), her T4 score returned to 26, with the participant attributing the deterioration in her questionnaire to interpersonal events occurring after the treatment window rather than to a recurrence of the original presenting problem. Consistent with the trial’s standing intention-to-treat policy, R59 is retained in the primary durability estimate, with a sensitivity row excluding her reported alongside; her case is discussed in Section 4.4 in relation to the personality-maturity construct carried forward to RCT #2. Durability and waitlist comparisons are tabulated in Table 5.

**Table 5.**
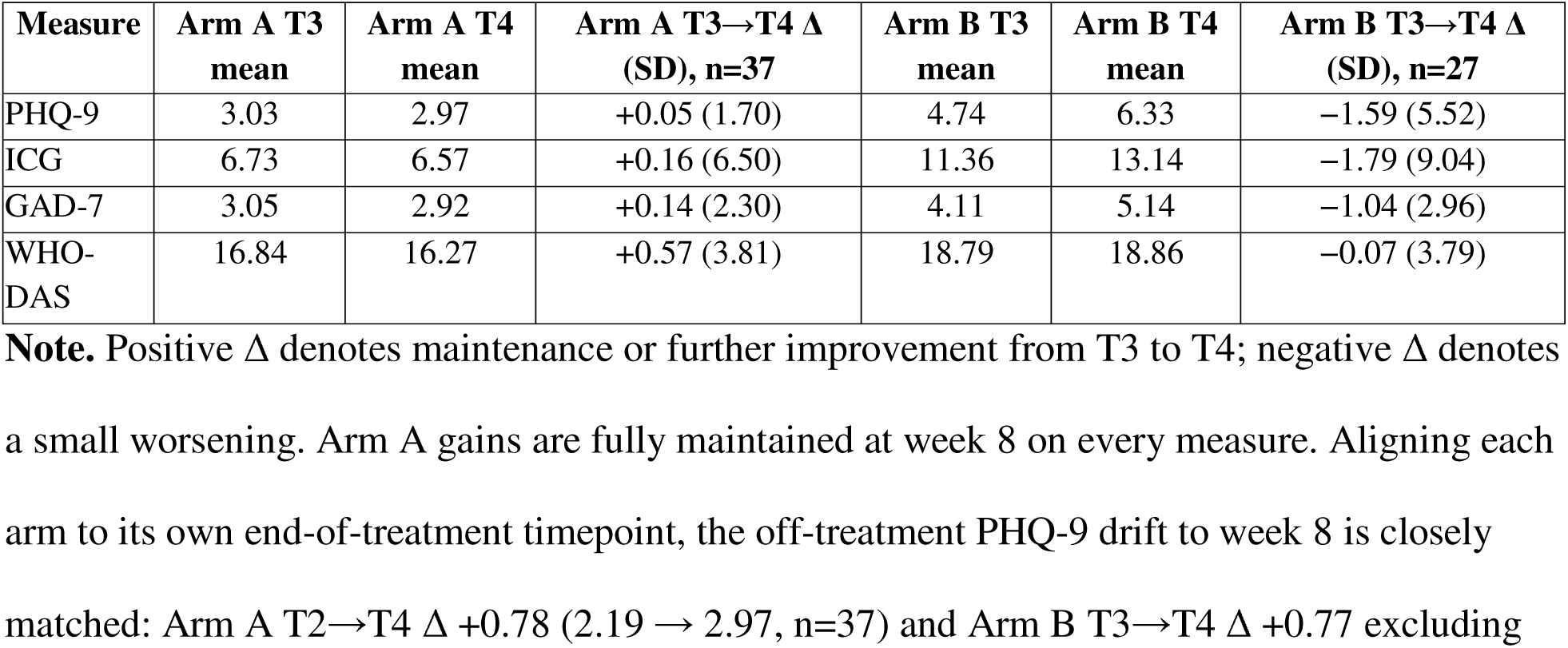
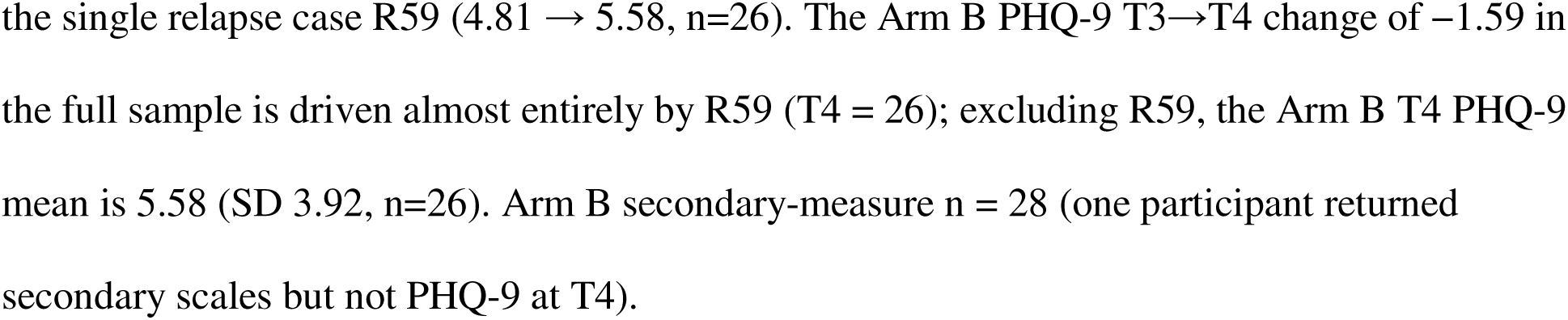
Durability of treatment gains at the T4 (week-8) follow-up, by arm.

**Table 5a.** Arm A early durability (T2 → T3) and Arm B waitlist window (T0 → T2).

| Measure | Arm A T2→T3 (n=37), $\Delta$ (SD) | Arm B T0→T2 (n=32), $\Delta$ (SD) |
| --- | --- | --- |
| PHQ-9 | -0.84 (2.34) | 0.22 (5.19) |
| ICG | 4.68 (9.96) | -2.44 (15.58) |
| GAD-7 | 0.84 (4.32) | -0.16 (3.27) |
| WHO-DAS | 1.35 (4.77) | -3.16 (8.17) |

Negative $\Delta$ values denote a worsening from the earlier timepoint to the later one.

### 3.7 Sensitivity analyses

Excluding the six Arm B participants whose T2 PHQ-9 fell below the protocol threshold (R16, R17, R32, R65, R68, R82) reproduced the eligible per-protocol Arm B result by construction (PHQ-9 T2 → T3 Δ 14.19, n=26), since all six failed the eligibility criterion built into the eligible-paired population. The eligible-paired Arm B population comprises all 26 participants with T2 PHQ-9 ≥ 9 and both T2 and T3 values observed, excluding only R68; it includes R84 (T2 = 20, T3 = 6), which a later coordinator count confirmed against the master file. One of the six sub-threshold cases, R68, reached PHQ-9 = 4 (remission) at the end of the waitlist window and was not advanced to active treatment; her T3 questionnaire was completed in error owing to a coordinator miscommunication and her T3 values are excluded from all outcome analyses. The single-exclusion sensitivity (R82 only, the sole sub-threshold participant who received treatment in error) shifted the Arm B T2 → T3 PHQ-9 reduction by less than 0.2 points; conclusions are unchanged.

Excluding the one Arm A late T2 submitter shifted the Arm A T0 → T2 PHQ-9 reduction by less than 0.1 PHQ-9 points; conclusions are unchanged. Excluding R69 (Arm B; deferred final session) shifted the Arm B T2 → T3 PHQ-9 reduction by less than 0.3 points; conclusions are unchanged. One Arm B participant submitted her T3 questionnaire five days after the deadline; consistent with the SAP standing late-submission policy, her data are retained in the ITT population, and a sensitivity row excluding this submission shifts the Arm B T2 → T3 PHQ-9 reduction by less than 0.3 points. Sensitivity analysis details are tabulated in Table 6.

**Table 6.**
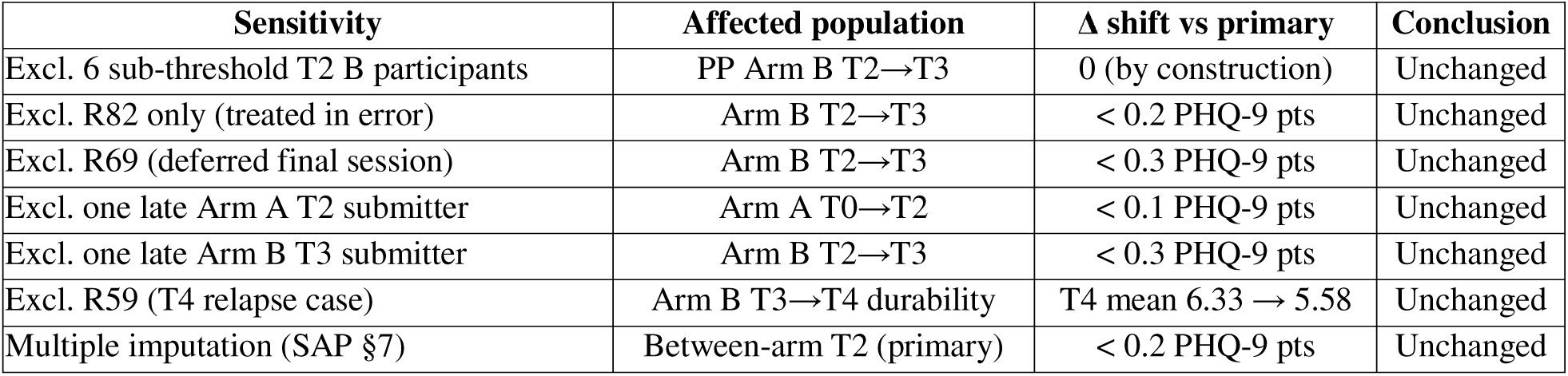
Sensitivity analyses on the primary outcome.

### 3.8 Exploratory moderator analyses

Baseline covariates were examined for association with PHQ-9 change (T0 → T2) in Arm A (n=38 completers). The Working Alliance Inventory-Short Revised (WAI-SR) showed near-zero correlation with outcome (r = −0.18). Credibility/Expectancy Questionnaire (CEQ) showed r = −0.05. General Self-Efficacy Scale (GSE) showed r = −0.10. All four custom readiness items showed |r| < 0.31. Age (r = −0.04) and sex (r = 0.12) showed near-zero correlations. These null findings suggest that the treatment effect is not attributable to pre-existing therapeutic alliance, treatment expectancy, self-efficacy, or a specific demographic subgroup.

### 3.9 Therapist experience and outcome

An exploratory analysis examined the association between therapist lifetime session count and mean PHQ-9 change across both arms (N=64 treated participants, 6 therapists). Mean PHQ-9 change increased monotonically with therapist experience: T1 (95 lifetime sessions, n=2) Δ 9.0; T2 (1,107 sessions, n=14) Δ 12.2; T3 (1,314 sessions, n=10) Δ 14.3; T4 (2,331 sessions, n=15) Δ 13.5; T5 (3,520 sessions, n=16) Δ 16.4; T6 (3,657 sessions, n=7) Δ 18.6. The participant-weighted Pearson correlation was r = 0.89 (unweighted r = 0.93). A participant-level analysis (N=64) yielded a Pearson correlation of r = 0.31 (p ≈ 0.012) between therapist lifetime session count and individual PHQ-9 change, confirming that the ecological association is not an artefact of grouped data. This finding should be interpreted with caution given the small number of therapists (N=6 ecological units), but the monotonic dose-response pattern is difficult to attribute to ceiling artefact or measurement bias.

### 3.10 Clinical threshold analysis

At baseline, no participant in either arm met PHQ-9 remission criteria (< 5), and 93% of Arm A and 88% of Arm B scored ≥ 10 (moderate-to-severe depression). At T2, 30 of 38 Arm A completers (79%) had achieved remission (PHQ-9 < 5) and none scored ≥ 10; in Arm B, 1 of 32 (3%) was in remission and 25 (78%) remained ≥ 10. At T3, 28 of 37 Arm A completers (76%) maintained remission, 3 (8%) had returned above PHQ-9 = 10; among Arm B post-crossover completers, 12 of 26 (46%) achieved remission and 1 (4%) remained ≥ 10. At T4, 25 of 37 Arm A completers (68%) remained in remission and 11 of 27 Arm B completers (41%) were in remission.

### 3.11 Deterioration and safety

Per the Statistical Analysis Plan (§14), symptomatic deterioration triggering an early coordinator-initiated session was pre-defined as a PHQ-9 increase of 5 or more points or a PHQ-9 item-9 (suicidal ideation) increase of 2 or more points, evaluated at the T1 early-escalation timepoint relative to baseline, as the protocol uses T1 for the early-session-escalation decision. Applying this criterion to the participants with a T1 measurement, two Arm B participants met the 5-point threshold during the waitlist window: R6 (T0 18 → T1 25, +7) and R12 (T0 18 → T1 23, +5). Both were offered an urgent coordinator-initiated session and completed the subsequent assessment before therapy began, so that the pre-treatment baseline was preserved; both then entered crossover treatment and showed substantial post-crossover PHQ-9 reductions. No participant met the item-9 escalation criterion at any timepoint, and no serious adverse events, hospitalisations, or safety referrals beyond the coordinator-initiated urgent-session offer were recorded. The single post-treatment relapse at T4 (R59, described in Section 3.6) emerged after treatment rather than during the waitlist window and did not involve emergent suicidal ideation. Waitlist-window deterioration of this kind is an expected feature of an untreated control arm rather than a treatment-emergent harm, and the deterioration log is tabulated in Table 7.

**Table 7.** Symptomatic deterioration during the Arm B waitlist window, evaluated at T1 (SAP §14 early-escalation threshold: PHQ-9 increase ≥ 5 points or item-9 increase ≥ 2 points, relative to baseline).

| Participant | PHQ-9 T0 | PHQ-9 T1 | Increase | Item-9 escalation | Action taken | Post-crossover outcome |
| --- | --- | --- | --- | --- | --- | --- |
| R6 | 18 | 25 | +7 | No | Urgent session offered; baseline captured pre-therapy | Reduced post-crossover |
| R12 | 18 | 23 | +5 | No | Urgent session offered; baseline | Reduced post- |

| Participant | PHQ-9<br>T0 | PHQ-9<br>T1 | Increase | Item-9<br>escalation | Action taken | Post-crossover<br>outcome |
| --- | --- | --- | --- | --- | --- | --- |
|  |  |  |  |  | captured pre-therapy | crossover |

**Note.** The deterioration criterion governing early coordinator-initiated escalation is evaluated at T1, the protocol-specified early-escalation timepoint, relative to baseline. Two Arm B participants with a T1 measurement crossed the 5-point threshold during the untreated waitlist window (R6, +7; R12, +5), matching the contemporaneous coordinator escalation log. No participant met the item-9 (suicidal ideation) escalation criterion at any timepoint, and no serious adverse events occurred. Waitlist-window symptom increase is an expected feature of an untreated control condition; both participants then entered crossover treatment and showed substantial PHQ-9 reductions.

### 3.12 Exploratory within-session mechanism checklist

The protocol pre-specified a within-session change-mechanism checklist in which Arm A participants recorded, after each session, how many of eleven defined change mechanisms they had identified. The measure was completed by a subset of Arm A participants (21 at session 1, 28 at session 2, 23 at session 3, 3 at session 4), and the data are therefore exploratory and cannot support formal mediation analysis. Two descriptive patterns are noted. The mean number of mechanisms identified rose across sessions (6.29 at session 1, 7.71 at session 2, 8.57 at session 3), consistent with progressive within-treatment insight. The number of mechanisms identified at session 1 showed a positive but modest association with subsequent PHQ-9 reduction (Pearson r = 0.31, n = 21). Both observations are hypothesis-generating only, are reported as a pre-specified exploratory analysis, and are interpreted in the Discussion (Section 4.3) rather than as confirmatory results.

## 4. Discussion

### 4.1 Principal findings

In a single-site randomised controlled trial of 84 adults with PHQ-9 ≥ 9 and active psychological distress related to adverse life events, Painhunting therapy produced a 14.25-point reduction in PHQ-9 at two weeks post-randomisation relative to a waitlist control (Cohen d = 2.78, p < 0.001). Within-arm paired analyses across each arm’s active-treatment window showed concordant reductions in both arms (Δ 14.71 in Arm A T0→T2 versus Δ 14.19 in Arm B T2→T3). Two findings carry particular weight. First, the between-arm magnitude was replicated within the trial when the waitlist arm itself crossed over to treatment, on a separate subgroup in a separate calendar window. Second, the gains were durable at the week-8 follow-up, and the durability behaved almost identically across arms once each is aligned to its own end-of-treatment timepoint: the off-treatment drift to week 8 was 0.78 PHQ-9 points in Arm A (T2 → T4) and 0.77 points in Arm B (T3 → T4, excluding a single documented relapse, R59), a small rebound of under one point from treated nadirs of 2 to 5 against baselines of 17 to 18. This rebound then stabilised rather than continuing downward, shown directly by the immediate-treatment arm being essentially unchanged from four to eight weeks (PHQ-9 Δ +0.05), with concordant maintenance on grief, anxiety, and disability measures and 68% of the arm in remission one month off-treatment. Durability at this interval distinguishes a genuine therapeutic change from a transient immediate-post-session improvement and is the property most relevant to the clinical value of a brief three-to-four-session intervention. Secondary measures (ICG, GAD-7, WHO-DAS) moved in the same direction and to comparable magnitude at every timepoint. The waitlist window in Arm B confirmed symptom stability in the absence of treatment (PHQ-9 Δ 0.22, p = 0.81), supporting a treatment-attributable interpretation of the Arm A reduction.

### 4.2 Comparison to the literature

The observed between-arm effect (Cohen d = 2.78) sits above the range typically reported in waitlist-controlled trials of brief structured psychotherapies for depression. Cuijpers et al. (2018) reported a pooled d of 0.72 across 385 psychotherapy comparisons. Patel et al. (2017) reported d = 0.57 for the Healthy Activity Programme, a lay-delivered behavioural activation intervention in India. Bolton et al. (2003) reported d = 1.42 for group interpersonal therapy in rural Uganda, one of the largest effects in the task-shifting literature. Richards et al. (2016) reported d = 0.95 for collaborative care versus usual care in primary-care depression.

Four features of the present trial argue that the observed magnitude, while large, reflects a genuine treatment signal rather than methodological artefact. First, the Arm B crossover replication (Δ 14.19) matches the Arm A treatment-window reduction (Δ 14.71) to within half a PHQ-9 point, on a separate subgroup treated in a separate calendar window. A chance result would not replicate at this precision. Second, the Arm B waitlist window is flat (Δ 0.22, p = 0.81), ruling out time effects and regression to the mean as explanations for the Arm A reduction. Third, all four outcome instruments (PHQ-9, ICG, GAD-7, WHO-DAS) move concordantly in both arms, a pattern inconsistent with single-instrument measurement bias. Fourth, baseline covariates hypothesised to inflate treatment effects (therapeutic alliance [WAI-SR, r = −0.18], treatment expectancy [CEQ, r = −0.05], self-efficacy [GSE, r = −0.10], age [r = −0.04], and sex [r = 0.12]) showed near-zero associations with PHQ-9 change in Arm A, arguing against expectancy effects, allegiance bias, or demographic selection as primary drivers.

A structural factor that may contribute to the observed magnitude is session duration. Painhunting sessions run 90 to 120 minutes, approximately twice the standard 50-minute CBT session. This dosing asymmetry is consistent with dose-response relationships documented in the broader psychotherapy literature and should be considered when comparing the present effect to trials using shorter sessions. The planned RCT #2 will permit a direct comparison against manualised CBT, which will yield a more interpretable relative effect.

### 4.3 Strengths

Pre-registration of the protocol and the statistical analysis plan, with the SAP locked prior to dataset lock, constrains analytical degrees of freedom. The crossover design provides a within-trial replication: the Arm B post-crossover reduction reproduces the Arm A treatment-window reduction to within 0.5 PHQ-9 points, a stronger evidentiary position than either arm in isolation. The four-measure outcome panel (depression, complicated grief, anxiety, functional disability) captures multiple domains rather than a single screening instrument. The therapist roster of six delivered sessions in varying counts, and the exploratory therapist-experience analysis (Section 3.9) shows a monotonic dose-response relationship that is consistent with protocol fidelity rather than therapist-specific idiosyncrasy.

The null associations between baseline covariates (WAI-SR, CEQ, GSE, age, sex) and PHQ-9 change (Section 3.8) address a common critique of developer-led trials. The treatment effect does not appear to be carried by participants with unusually high therapeutic alliance expectations, high treatment credibility, or a specific demographic profile, which reduces the plausibility of allegiance bias as the primary driver of the observed effect.

All six therapists who delivered the intervention have non-psychology professional backgrounds; none holds a psychology degree. The intervention was learned over approximately six months of structured certification. Even the least experienced therapist in the trial (95 lifetime sessions) produced a mean PHQ-9 reduction of 9.0 points, above the conventional threshold for a clinically meaningful change. This pattern is consistent with the WHO task-shifting model for mental health interventions in low-resource settings, as exemplified by mhGAP, the Friendship Bench, and the Healthy Activity Programme, and suggests that the intervention protocol can be delivered effectively by non-specialist providers after structured training.

The pre-specified within-session mechanism checklist (Section 3.12) offers a tentative, hypothesis-generating window onto the intervention’s proposed mechanism. The number of change mechanisms participants identified rose across successive sessions (6.29, 7.71, 8.57), and the count reported after the first session was modestly associated with subsequent depression reduction (r = 0.31). These patterns are consistent with the theorised account in which therapeutic benefit accrues through progressive recognition and processing of event-linked change mechanisms. They must be read with considerable caution: the checklist was completed by only half to two-thirds of the Arm A participants, the analysis is correlational, and no formal mediation model is supportable on these data. We report the patterns because the measure was pre-specified in the protocol and because they identify a concrete and testable mediational hypothesis for the planned RCT #2, in which the checklist will be administered as a complete-data instrument with a pre-registered mediation analysis.

### 4.4 Limitations

Differential retention is the principal limitation. Arm B retention at T3 was 64% versus 88% in Arm A, a 24-percentage-point gap that is itself an outcome of the waitlist design but one that raises the prospect of a selection effect in the per-crossover Arm B population. We addressed this by reporting the eligible-paired n for each within-arm contrast and by pre-specifying per-protocol and sensitivity populations. Outcome assessment was self-report via Google Forms in a setting in which a Russian-language coordinator was available to clarify item wording; while the coordinator was instructed not to influence response content, we cannot fully exclude unblinded contact effects. The trial is single-site and predominantly female (88% across both arms), constraining generalisability to other settings and to male presentations of event-related depression. Six Arm B participants with T2 PHQ-9 below the protocol threshold were not flagged contemporaneously, and one (R82) was treated in error; sensitivity analyses excluding these participants did not change conclusions, but the operational disclosure is a real protocol deviation that constrains some between-arm interpretations.

An optional Painhunting adherence audit was not funded for this trial, meaning that we cannot independently verify treatment fidelity on the Painhunting arm. The CTSR-rated CBT adherence audit planned for RCT #2 creates an asymmetry that the present trial’s design cannot resolve.

Several participants’ post-treatment trajectories warrant comment as hypothesis-generating clinical observations. Two Arm A participants (R28, R58) with elevated T3 PHQ-9 scores (14 and 10, respectively) shared a common profile: both scored in the lowest quartile of the team’s internal personality-maturity assessment. A third participant (R3, T3 PHQ-9 = 11) had an active, unresolved divorce stressor that continued to produce session-to-session instability independent of the grief-processing work. The extended follow-up added a fourth and particularly clear case: one Arm B participant (R59) reduced from a crossover-window PHQ-9 of 22 to 3 immediately post-treatment, then relapsed to 26 at T4, attributing the deterioration in her own questionnaire to disillusioning interpersonal events that arose after treatment rather than to a return of the original presenting complaint. The treating team characterised her as combining early bereavement, recent spousal loss, and a marked tendency toward idealisation followed by abrupt disillusionment, a profile they place at the low end of the same personality-maturity construct. These cases jointly suggest two distinct predictors of post-treatment instability: low personality maturity and the presence of an active, unresolved stressor at follow-up. Neither predictor is testable in the present trial (the maturity instrument is unvalidated, and the case count is statistically inadequate), but both are incorporated into the pre-registered moderator panel for RCT #2 as the personality-maturity scale (pending psychometric validation in RCT #2) and an active-stressor screening item. The R59 trajectory also motivates the separate longer-term treatment protocol under development for participants whose vulnerability to post-treatment relapse reflects this maturity dimension rather than the adequacy of the index intervention.

The present trial was designed and led by the intervention developer, introducing the potential for allegiance bias in treatment delivery and outcome interpretation. Although baseline covariate analyses showed near-zero associations with outcome (Section 3.8), independent replication by investigator teams without developer involvement remains necessary to establish generalisability of the observed effect sizes. Protocol manualisation and therapist certification standards are being developed to support investigator-independent replication.

The trial was conducted without external funding, which constrained two design parameters. First, the session protocol was limited to three to six sessions; a funded design would have permitted continuation to full remission for participants above threshold at session three. Second, the crossover eligibility threshold was set at PHQ-9 ≥ 9 rather than the more stringent remission criterion of PHQ-9 < 5, again to limit therapist burden. Both constraints represent a conservative bias: the observed effect sizes likely underestimate the full therapeutic potential of the intervention under adequately resourced conditions.

Session-level PHQ-9 data, available for a subset of participants, suggested progressive within-treatment reductions from the first session onward; formal dose-response analysis is planned for a dedicated report.

### 4.5 Implications and next steps

The trial establishes initial evidence that Painhunting therapy produces large and durable reductions in depressive, grief, anxiety, and disability outcomes relative to waitlist control. The next step is an active-comparator trial against manualised CBT, which is the design of the planned RCT #2 (Painhunting versus CBT for Event-Related Depression). RCT #2 will preserve the methodological architecture of the present trial (stratified randomisation, blinded outcome assessment, SAP-locked analysis populations) and will add a pre-registered moderator panel including a personality-maturity scale (pending psychometric validation in RCT #2) and an active-stressor screening item, both of which arose from the clinical observations in the present trial. The nested therapist-effects analysis planned for RCT #2 will formally test the exploratory therapist-experience association observed here.

### 4.6 Conclusion

Painhunting therapy, a brief structured psychotherapeutic intervention of three to four sessions developed and delivered in Kazakhstan, produced a 14.25-point reduction in PHQ-9 at two weeks post-randomisation relative to waitlist control (Cohen d = 2.78, p < 0.001), with within-trial replication during Arm B crossover and durability of gains at the week-8 follow-up, approximately one month off-treatment, where the immediate-treatment arm was essentially unchanged from its post-treatment level and two-thirds of participants remained in remission. Findings support proceeding to a confirmatory active-comparator trial against manualised CBT.

## Acknowledgements

The authors thank all 84 trial participants for their time and commitment. We thank the trial coordinators, study administrator, blinded assessors, and the six certified Painhunting therapists who delivered the intervention for their contributions to the study. Ethics oversight was provided by the al-Farabi KazNU Local Ethics Committee (IRB00010790).

## Author Contributions (CRediT)

O.S.: conceptualisation, methodology, investigation, data curation, project administration, and writing (review and editing). U.S.: investigation, supervision, validation, and writing (review and editing). V.B.: methodology, formal analysis, software, data curation, visualisation, writing (original draft), and writing (review and editing).

## Data Availability

The analysis dataset, de-identified data dictionary, statistical analysis plan, and analysis code are available on the Open Science Framework at https://osf.io/s8h74/. The trial is registered on ClinicalTrials.gov (NCT07490691).

Trial registration: ClinicalTrials.gov NCT07490691 (status: Completed; primary completion date 6 June 2026). Ethics approval: al-Farabi KazNU LEC (IRB00010790), IRB-1970. Version 1.7, dated 8 June 2026. Manuscript draft prepared against the master file dated 6 June 2026 (Final Master file for Vafa 6 Jun 26 with T4.xlsx), which incorporates the complete T4 (week-8) follow-up dataset.

