## Supplemental Tables for "Efficacy of Painhunting Therapy for Event-Related Depression: A Randomized Controlled Trial with Crossover Replication"

**Painhunting Therapy RCT: Tables (Manuscript v1.7, T4)**

*Companion to Painhunting_RCT_T4_Manuscript_v1.7_2026-06-08*

**Table 1. Baseline characteristics by arm.**

| **Characteristic** | **Arm A (n=42)** | **Arm B (n=42)** |
| --- | --- | --- |
| Age, years (mean ± SD) | 41.6 ± 10.1 | 42.5 ± 9.2 |
| Female sex, n (%) | 36 (86%) | 39 (93%) |
| PHQ-9 (mean ± SD) | 17.31 ± 5.14 | 16.69 ± 4.92 |
| ICG (mean ± SD) | 41.57 ± 19.48 | 36.02 ± 17.16 |
| GAD-7 (mean ± SD) | 13.74 ± 5.68 | 11.90 ± 6.04 |
| WHO-DAS (mean ± SD) | 30.79 ± 10.24 | 27.60 ± 10.05 |
| LTE events, mean | 3 | 2 |
| GSE (mean ± SD) | 25.0 ± 6.5 | 26.7 ± 5.6 |
| WAI-SR (mean ± SD) | 77.4 ± 15.0 | 76.6 ± 12.4 |
| CEQ (mean ± SD) | 7.8 ± 1.7 | 7.7 ± 1.7 |
| Stratum BD, n | 22 | 22 |
| Stratum AD, n | 13 | 14 |
| Stratum BC, n | 5 | 5 |
| Stratum AC, n | 2 | 1 |

*Note. PHQ-9, Patient Health Questionnaire-9; ICG, Inventory of Complicated Grief; GAD-7, Generalised Anxiety Disorder-7; WHO-DAS, WHO Disability Assessment Schedule; LTE, List of Threatening Experiences; GSE, General Self-Efficacy; WAI-SR, Working Alliance Inventory-Short Revised; CEQ, Credibility/Expectancy Questionnaire. Values are mean ± SD unless noted.*

**Table 2. Participant retention and assessment completion by timepoint.**

| **Timepoint** | **Arm A completed, n (%)** | **Arm B completed, n (%)** |
| --- | --- | --- |
| T0 (baseline) | 42 (100%) | 42 (100%) |
| T2 (2 weeks) | 38 (90%) | 32 (76%) |
| T3 (4 weeks) | 37 (88%) | 27 (64%) |
| T4 (8 weeks) | 37 (88%) | 27 (64%) |

*Note. Percentages are of the n=42 randomised per arm. Completion reflects assessment submission at each timepoint. Arm B T4 secondary-measure completion was 28 (one participant returned secondary scales but not the PHQ-9 at T4).*

**Table 3. Primary outcome: between-arm contrast at T2 (PHQ-9).**

| **Population** | **Arm A mean (SD)** | **Arm B mean (SD)** | **Difference** | **Cohen d (95% CI)** | **p** |
| --- | --- | --- | --- | --- | --- |
| ITT (n A=38, B=32) | 2.32 (2.59) | 16.56 (6.76) | −14.25 | 2.78 (2.19–3.76) | < 0.001 |
| PP (excl. 6 sub-threshold B; n B=26) | 2.32 (2.59) | 18.92 (5.04) | −16.61 | 4.15 (3.38–5.53) | < 0.001 |
| Sensitivity (excl. R82 only; n B=31) | 2.32 (2.59) | 16.84 (6.69) | −14.52 | 2.86 (2.25–3.93) | < 0.001 |

*Note. Primary contrast is PHQ-9 at T2. SD values are raw group descriptive standard deviations and Cohen d is the between-arm standardised mean difference computed with the pooled raw SD (square root of the mean of the two group variances), so that d reproduces directly from the reported means and SDs. The ITT row contrasts all T2 completers (Arm A n=38, Arm B n=32); the PP row excludes the six Arm B participants below the PHQ-9 ≥ 9 protocol threshold at T2 (Arm B n=26) against the same Arm A completers. Confidence intervals are nonparametric bootstrap (10,000 resamples, percentile method). Values reconciled against the master file dated 6 June 2026.*

**Table 4. Within-arm treatment effects (Morris d): Arm A T0→T2 and Arm B post-crossover T2→T3.**

| **Measure** | **Arm A T0→T2 (n=38), Δ (SD)** | **Morris d (95% CI)** | **Arm B T2→T3 (n=26), Δ (SD)** | **Morris d (95% CI)** |
| --- | --- | --- | --- | --- |
| PHQ-9 | 14.71 (5.90) | 2.80 (2.41–3.46) | 14.19 (5.97) | 2.77 (2.11–4.00) |
| ICG | 29.74 (22.21) | 1.50 (1.17–1.96) | 32.50 (16.20) | 1.93 (1.35–3.32) |
| GAD-7 | 9.45 (7.15) | 1.66 (1.19–2.45) | 9.88 (5.74) | 1.85 (1.36–2.69) |
| WHO-DAS | 12.42 (10.74) | 1.19 (0.87–1.66) | 16.92 (11.12) | 1.69 (1.28–2.42) |

*Note. Morris d (pretest–posttest control design) reported within arm. Arm B values are post-crossover (T2→T3).*

**Table 5. Durability of treatment gains at the T4 (week-8) follow-up, by arm.**

| **Measure** | **Arm A T3 mean** | **Arm A T4 mean** | **Arm A T3→T4 Δ (SD), n=37** | **Arm B T3 mean** | **Arm B T4 mean** | **Arm B T3→T4 Δ (SD), n=27** |
| --- | --- | --- | --- | --- | --- | --- |
| PHQ-9 | 3.03 | 2.97 | +0.05 (1.70) | 4.74 | 6.33 | −1.59 (5.52) |
| ICG | 6.73 | 6.57 | +0.16 (6.50) | 11.36 | 13.14 | −1.79 (9.04) |
| GAD-7 | 3.05 | 2.92 | +0.14 (2.30) | 4.11 | 5.14 | −1.04 (2.96) |
| WHO-DAS | 16.84 | 16.27 | +0.57 (3.81) | 18.79 | 18.86 | −0.07 (3.79) |

*Note. Positive Δ denotes maintenance or further improvement from T3 to T4; negative Δ denotes a small worsening. Arm A gains are fully maintained at week 8 on every measure. Aligning each arm to its own end-of-treatment timepoint, the off-treatment PHQ-9 drift to week 8 is closely matched: Arm A T2→T4 Δ +0.78 (2.19 → 2.97, n=37) and Arm B T3→T4 Δ +0.77 excluding the single relapse case R59 (4.81 → 5.58, n=26). The Arm B PHQ-9 T3→T4 change of −1.59 in the full sample is driven almost entirely by R59 (T4 = 26). Arm B secondary-measure n = 28.*

**Table 5a. Early durability (Arm A T2→T3) and waitlist window (Arm B T0→T2).**

| **Measure** | **Arm A T2→T3 (n=37), Δ (SD)** | **Arm B T0→T2 (n=32), Δ (SD)** |
| --- | --- | --- |
| PHQ-9 | −0.84 (2.34) | 0.22 (5.19) |
| ICG | 4.68 (9.96) | −2.44 (15.58) |
| GAD-7 | 0.84 (4.32) | −0.16 (3.27) |
| WHO-DAS | 1.35 (4.77) | −3.16 (8.17) |

*Note. Arm A T2→T3 captures early off-treatment durability (2 weeks off-treatment); negative Δ denotes mild regression toward baseline. Arm B T0→T2 is the untreated waitlist window.*

**Table 6. Sensitivity analyses.**

| **Sensitivity** | **Affected population** | **Δ shift vs primary** | **Conclusion** |
| --- | --- | --- | --- |
| Excl. 6 sub-threshold T2 B participants | PP Arm B T2→T3 | 0 (by construction) | Unchanged |
| Excl. R82 only (treated in error) | Arm B T2→T3 | < 0.2 PHQ-9 pts | Unchanged |
| Excl. R69 (deferred final session) | Arm B T2→T3 | < 0.3 PHQ-9 pts | Unchanged |
| Excl. one late Arm A T2 submitter | Arm A T0→T2 | < 0.1 PHQ-9 pts | Unchanged |
| Excl. one late Arm B T3 submitter | Arm B T2→T3 | < 0.3 PHQ-9 pts | Unchanged |
| Excl. R59 (T4 relapse case) | Arm B T3→T4 durability | T4 mean 6.33 → 5.58 | Unchanged |
| Multiple imputation (SAP §7) | Between-arm T2 (primary) | < 0.2 PHQ-9 pts | Unchanged |

*Note. All pre-specified and post-hoc sensitivity analyses, including exclusion of the single T4 relapse case (R59) and the SAP §7 multiple-imputation analysis on the primary contrast, leave the primary conclusion unchanged.*

**Table 7. Symptomatic deterioration during the Arm B waitlist window, evaluated at T1 (SAP §14).**

| **Participant** | **PHQ-9 T0** | **PHQ-9 T1** | **Increase** | **Item-9 escalation** | **Action taken** | **Post-crossover outcome** |
| --- | --- | --- | --- | --- | --- | --- |
| R6 | 18 | 25 | +7 | No | Urgent session offered; baseline captured pre-therapy | Reduced post-crossover |
| R12 | 18 | 23 | +5 | No | Urgent session offered; baseline captured pre-therapy | Reduced post-crossover |

*Note. The early-escalation criterion (SAP §14) is a PHQ-9 increase ≥ 5 points or an item-9 (suicidal ideation) increase ≥ 2 points, evaluated at T1 relative to baseline, as the protocol uses T1 for the early-session-escalation decision. Two Arm B participants with a T1 measurement crossed the 5-point threshold during the untreated waitlist window (R6, +7; R12, +5), matching the contemporaneous coordinator escalation log; none met the item-9 criterion and no serious adverse events occurred.*
